# Deep Learning in Automating Breast Cancer Diagnosis from Microscopy Images

**DOI:** 10.1101/2023.06.15.23291437

**Authors:** Qiangqiang Gu, Naresh Prodduturi, Steven N. Hart

## Abstract

**Context:** Breast cancer is one of the most common cancers in women. With early diagnosis, some breast cancers are highly curable. However, the concordance rate of breast cancer diagnosis from histology slides by pathologists is unacceptably low. Classifying normal versus tumor breast tissues from microscopy images of breast histology is an ideal case to use for deep learning and could help to more reproducibly diagnose breast cancer. Since data preprocessing and hyperparameter configurations have impacts on breast cancer classification accuracies of deep learning models, training a deep learning classifier with appropriate data preprocessing approaches and optimized hyperparameter configurations could improve breast cancer classification accuracy.

**Methods and Material:** Using 12 combinations of deep learning model architectures (i.e., including 5 non-specialized and 7 digital pathology-specialized model architectures), image data preprocessing, and hyperparameter configurations, the validation accuracy of tumor versus normal classification were calculated using the **B**re**A**st **C**ancer **H**istology (BACH) dataset.

**Results:** The DenseNet201, a non-specialized model architecture, with transfer learning approach achieved 98.61% validation accuracy compared to only 64.00% for the digital pathology-specialized model architecture.

**Conclusions:** The combination of image data preprocessing approaches and hyperparameter configurations have a profound impact on the performance of deep neural networks for image classification. To identify a well-performing deep neural network to classify tumor versus normal breast histology, researchers should not only focus on developing new models specifically for digital pathology, since hyperparameter tuning for existing deep neural networks in the computer vision field could also achieve a high (often better) prediction accuracy.

## 1. Introduction

Breast cancer is one of the leading cancer-related causes of death in women.^1^ Early-diagnosis for breast cancer can reduce the mortality rate for breast cancer patients given that 70-80% of patients with early-diagnosis of non-metastatic breast cancer are curable.^2^

Breast biopsy is the definitive way to diagnose breast cancer,^3^ however, the concordance rate between different pathologists in interpreting breast biopsies is relatively low (overall concordance rate is 75.3% with 48% concordance rate for atypia).^4^ To improve agreement, deep learning has shown success in solving broader computer vision problems,^5^ particularly in the medical image analysis field.^6^

The advent of whole slide imaging^7^ has heralded a new era in pathology research, enabling the detailed analysis of histological images through deep learning methodologies.^8^ This is highlighted in the work of Iizuka et al.,^9^ who successfully employed deep learning algorithms to identify gastric and colonic epithelial tumors within histological slide preparations. Their approach achieved remarkable levels of accuracy, as demonstrated by Area Under the Curve (AUC) values of 97% and 99% for the prediction of gastric adenocarcinoma and adenoma, respectively. Likewise, colonic adenocarcinoma and adenoma prediction achieved AUC values of 96% and 99%, respectively. These findings underscore the potential of deep learning-based image classifiers to enhance diagnostic precision, positioning it as a promising approach for distinguishing normal tissues from malignant neoplasms.

Differentiation of malignant tumors and normal tissues on histology slides can be achieved by two deep learning-based image classification approaches. First, non-specialized deep neural networks have been applied to group different classes of histology from microscopy images. Transfer learning^10^ is a popular non-specialized approach, which uses either the last layer or all layers of the pre-trained networks, including InceptionV3,^11^ DenseNet201,^12^ ResNet152,^13^ and VGG19^14^ models for image classification. One-shot learning,^15^ a distance-based classification model, is another non-specialized approach to predict the object categories from a few training samples. Koch, et al.^16^ adopted the one-shot learning model for image classification^17^ achieving near-state-of-the-art classification accuracy. Aside from general use networks, specialized deep neural networks have also been developed for microscopy images. The clustering-constrained attention multiple instance learning (CLAM) model^18^ is a digital pathology-specialized multi-class image classifier. CLAM is an attention-based weakly-supervised learning model that does not require large amounts of well-annotated training samples. CLAM is a unique approach in digital pathology, that ranks the patch-level feature importance by attention scores, then ranks information to train the final classifier.

Different deep learning models could affect the classification performance. However, hyper-parameter configurations^19^ and data preprocessing^20^ also have impacts on the performance of image classifiers. Zhou et al.^21^ proposed a comparative experiment to study the impacts of hyperparameters on deep learning model performance. They found the classification precision scores varied from 84.8% to 99.5% for a number of 36 combinations of deep convolutional neural networks (DCNN)-based a roadway crack classification problem. They tested various hyperparameter configurations, including learning rate, dropout, and batch size on 10,000 test images from laser-scanned roadway range image dataset (LRRD).^22^ In addition, Heidari et al.^23^ proposed a study to compare the performance of VGG16-based transfer learning approach with or without image preprocessing in classifying COVID-19, non-COVID-19 pneumonia, and non-pneumonia cases from 8,504 2D X-ray images. The authors yielded a 7.4% increase in overall classification accuracy of the VGG16-based classifier with image preprocessing compared with the model without pre-processing steps. This indicates that the image preprocessing could also alter the deep learning model performance. Therefore, the standard deep neural networks could achieve a better classification performance by hyperparameter tuning and selecting appropriate data pre-processing techniques. What is not known is how much of a difference hyperparameters, model architectures, or general versus domain specific architecture make on medically relevant images like those in digital pathology.

The **B**re**A**st **C**ancer **H**istology (BACH) dataset^24^ is a publicly available dataset of Hematoxylin and Eosin (H&E)-stained microscopy images of breast histology labeled into four classes (i.e., “normal”, “benign”, “*in situ* carcinoma” and “invasive carcinoma”). An ensemble network-based image classifier proposed by Marami et al.^25^ was the best performing model on the BACH dataset with the highest prediction accuracy. Their model was able to achieve an 84% accuracy for the four-class classification required by the BACH Challenge, but also achieved a 91.7% accuracy in classifying carcinoma versus non-carcinoma breast histology. The carcinoma versus non-carcinoma classification was made possible by using a binary classification model in which the images from “normal” and “benign” classes were reassigned into a single “non-carcinoma” class and images from “*in situ* carcinoma” and “invasive” carcinoma classes were reassigned into a single “carcinoma” class. However, the proposed approach by Marami et al. was to build a de novo algorithm using an ensemble of convolutional neural networks rather than fine tuning the conventional deep neural networks (i.e., ResNet,^13^ and InceptionResNet^26^). Therefore, the proposed study compared the performance of models with different combinations of hyperparameters and data pre-processing techniques, including custom versus purpose-built models.

## 2. Subjects and Methods

### 2.1. Data Preparation

Four hundred microscopy images of breast histology in ‘.tif’ format were downloaded from the BACH dataset. Out of the 400 images, there are 100 microscopy images from each of the “benign”, “normal”, “*in situ* carcinoma” and “invasive carcinoma” classes. To reorganize the images from the BACH dataset for binary carcinoma versus non-carcinoma classification, images in the “benign” or “normal” BACH classes are labeled the “non-carcinoma” class (i.e., class 0) and images within the “*in situ* carcinoma” or “invasive carcinoma” BACH classes are labeled the “carcinoma” class (i.e., class 1).

To create a 5-fold cross validation dataset, all 400 images were first randomly shuffled and divided into five groups. To maintain a balanced dataset in each of the five groups, each group ended up with 80 images, including 40 images each from the carcinoma and non-carcinoma classes. For each of the 5-folds, one of the five groups is selected as the validation set, while the remaining four groups are selected as the training set. The 5-fold cross validation dataset preparation was implemented using the Scikit-Learn Python package.^27^ Therefore, in each of the 5-folds, there are 320 images with 160 images from each of the carcinoma and non-carcinoma classes used for training, and 80 images with 40 images each from the carcinoma and non-carcinoma classes used for validation. Patches from 400 microscopy images were extracted and saved in the TFRecords file format with each TFRecords file including the image patch array, file name, width, and height of the image patch.^28^

CLAM required image patch-level feature vectors as the model training input data -rather than images - while the pre-trained InceptionV3, DenseNet201,^12^ ResNet152, VGG19, and one-shot learning model only required pixel data as input. **Sections 2.1.3 - 2.1.4** details the image feature extraction and normalization, specific for CLAM, while the **sections 2.1.1 - 2.1.2** describe patch extraction, image standardization, and scaling - all of which are identical for all deep neural networks.

Of note, it was also necessary to re-implement CLAM as it did not support the BACH files and some of the standardized profiling that are needed to perform. Comparing the re-implemented CLAM with the original source code confirmed there was no difference in classification outcomes. To make the comparison, A number of 40 H&E-stained malignant breast histology WSIs were downloaded from the Cancer Genome Atlas (TCGA) database.^29^ These 40 WSIs include 20 BRAF mutated and 20 wild-type malignant breast histology WSIs. Then, 10 cross-validation sets were created by randomly selecting 35 out of the total 40 WSIs for each of the 10 folds, and split into training, validation, and testing sets. In each cross-validation set, there were 15 WSIs in the training set, 10 WSIs in the validation set, and 10 other WSIs in the testing set. The extracted image patches were used to create the image feature vectors for all the WSIs in each of the 10 cross-validation sets without any image preprocessing.

#### 2.1.1. Image Patch Preparation

Each microscopy image from the BACH dataset has 2,048 × 1,536 × 3 pixels with a pixel scale of 0.42 *μm* × 0.42 *μm*.^24^ JPEG format images (n=19,200) of 256 × 256 × 3 pixels in were split into 5-fold cross-validation sets, with 15,360 image patches in the training (Class0: n=7,680; Class1=n=7,680) and 3,840 patches (Class0: n=1,920; Class1: n=1,920) in the validation (**Figure 1**).

**Figure 1.**
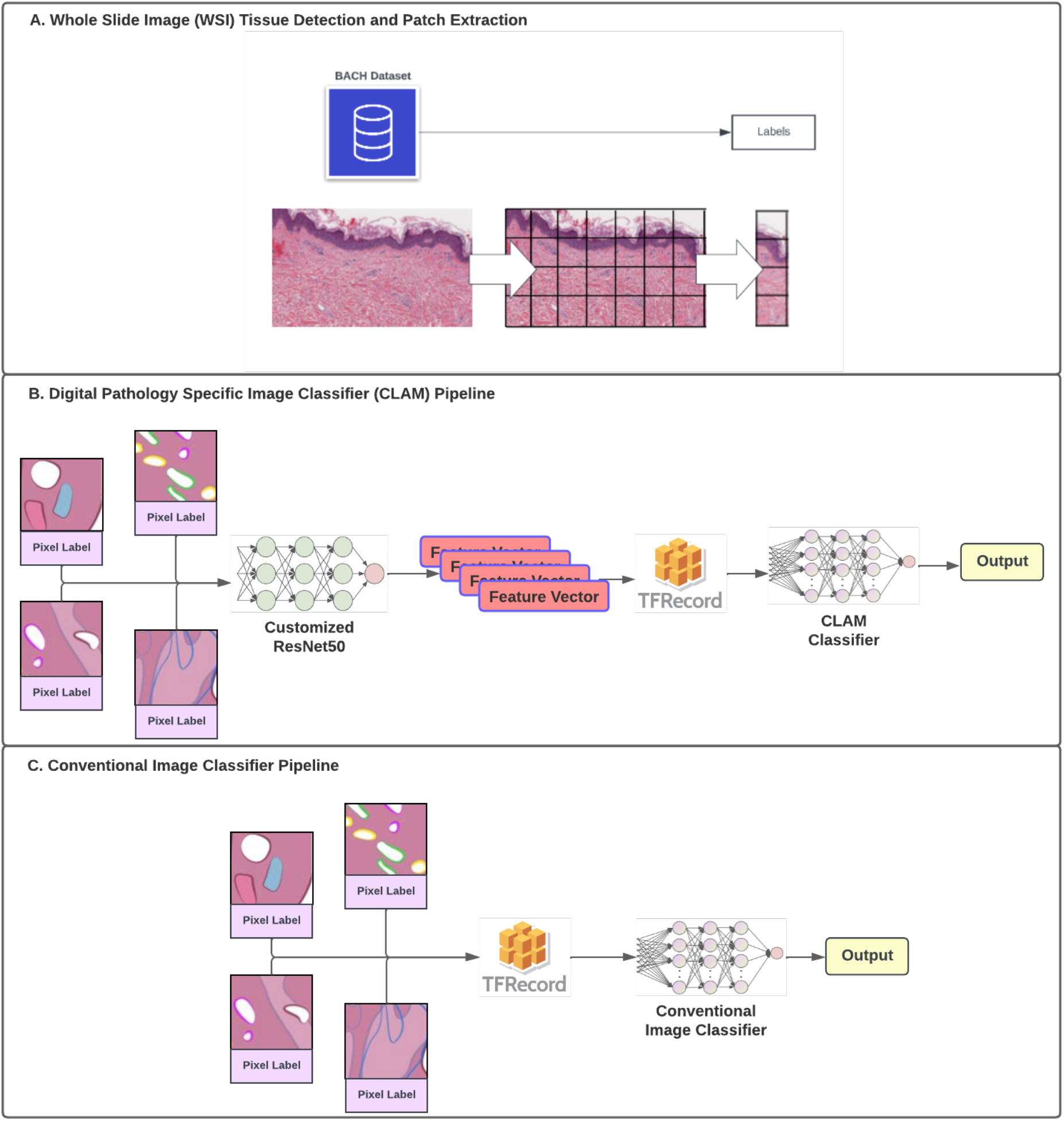
Pipeline Diagram for Digital Pathology-Specialized and Non-Specialized Image Classifiers. A). Whole Slide Image Tissue Detection and Patch Extraction; B). Digital Pathology-Specialized Image Classifier (CLAM) Pipeline; C). Non-Specialized Conventional Image Classifiers (i.e., DenseNet201, InceptionV3, One-Shot Learning, ResNet152, and VGG19) Pipeline.

#### 2.1.2. Image Standardization

Image standardization is an image rescaling technique that linearly scales each of the 3 RGB-channel (i.e., red, green, blue) image patches to a mean of 0 and variance of 1. The formula of this technique to compute the standardized image patch array 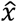 is:

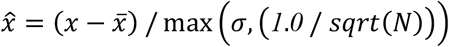

where,

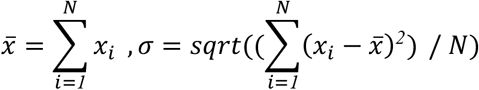

and *N* is denoted as the number of elements in each of the image patch *x*.

An additional image rescaling technique is also applied in one of the experiments in this study. The formula used to compute the rescaled image patch array 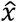 from the original image patch array *x* is:

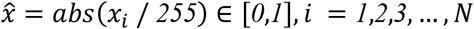

where *N* is denoted as the number of elements in each image patch *x*.

Details of the combinations of different image scaling methods and experiments were listed in the **Supplementary Table 1**.

#### 2.1.3. Image Feature Extraction

The pre-trained ResNet50 model on ImageNet^30^ was employed to extract image feature vectors for the preparation of CLAM model training. RGB channel image patches with dimensions of 256 × 256 × 3 were fed into this pre-trained ResNet50 model. Following processing through the third residual block of the pre-trained ResNet50 model, a 1,024-dimensional patch-level image feature vector was obtained (**Figure 1**).

#### 2.1.4. Image Feature Normalization

Image patch-level feature vectors are the required input for CLAM training. The L2 normalization^31^ was applied on the extracted 1,024-dimensional patch-level image feature vectors to generate the normalized patch-level image feature vectors.

Each of the L2 normalized patch-level 1,024-dimensional image feature vectors 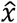 was computed from each of the original patch-level 1,024-dimensional image feature vectors *x* by the following,

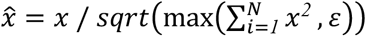

where *ε* has a default value of 1E-12, and *N* is denoted as the number of elements in each of the patch-level 1,024-dimensional image feature vectors *x*.

### 2.2. Model Training

#### 2.2.1. Transfer Learning with Pre-trained Deep Learning Models

Transfer learning was applied with different non-specialized model architectures, including InceptionV3, DenseNet201, ResNet152, and VGG19. These models were first pre-trained on ImageNet, then the last layer of these pre-trained models was trained on the H&E microscopy images from the BACH dataset. Training details of these models with the corresponding combinations of data preprocessing (i.e., image standardization, and image feature normalization), and hyperparameter configurations (i.e., learning rate, dropout rate, optimizers, loss functions, number of epochs, and batch size) are listed in **Supplementary Table 1**.

#### 2.2.2. One-Shot Learning

One-shot learning was applied to learn the domain features from microscopy images from the normal and tumor classes reorganized from the BACH dataset. This would have allowed the model to classify the normal versus tumor breast histology from microscopy images.

Training details of the combination of the one-shot learning model, image data preprocessing (i.e., image standardization, and image feature normalization), and hyperparameter configurations (i.e., learning rate, dropout rate, optimizers, loss functions, number of epochs, and batch size) are listed in **Supplementary Table 1**.

#### 2.2.3. Clustering-Constrained Attention Multiple Instance Learning (CLAM)

Microscopy images of breast histology from the BACH dataset are in ‘.tif’ format, which is not supported by the original CLAM implementation. A TensorFlow-version^28^ CLAM was re-implemented with three jointly trained neural networks (i.e., attention network,^32^ instance classifier, and bag classifier^33^). To ensure the re-implemented CLAM achieves a similar classification performance as the original CLAM, both the original and re-implemented CLAM were evaluated on 10 validation WSIs from each of the 10 cross-validation sets to compute the validation accuracy. Then a Student’s t-test ran on the AUC of the original and re-implemented CLAM on 10 validation WSIs from each of the 10 cross-validation sets to determine whether the re-implemented CLAM achieves a similar binary classification accuracy as the original CLAM.

Then, similar to the experiments that have been performed using the non-specialized classifiers as discussed on **section 2.2.1 - 2.2.2**, the validation accuracy of CLAM with 7 different combinations of data preprocessing (i.e., image standardization, and image feature normalization), and hyperparameter configurations (i.e., learning rate, dropout rate, optimizers, loss functions, number of epochs, and batch size) are listed in **Supplementary Table 1**.

All code, including the implementations of non-specialized and digital pathology-specialized model architectures, is publicly available at https://github.com/quincy-125/DP_BACH.

## 3. Results & Discussion

### 3.1. CLAM Reimplementation Results on TCGA Data

The AUC scores returned from both the original and re-implemented CLAM on 10 validation TCGA WSIs from each of the 10 cross-validation sets are shown in **Figure 2**. There was no significant difference between the performance of the original and re-implemented CLAM (p-value=0.67).

**Figure 2.**
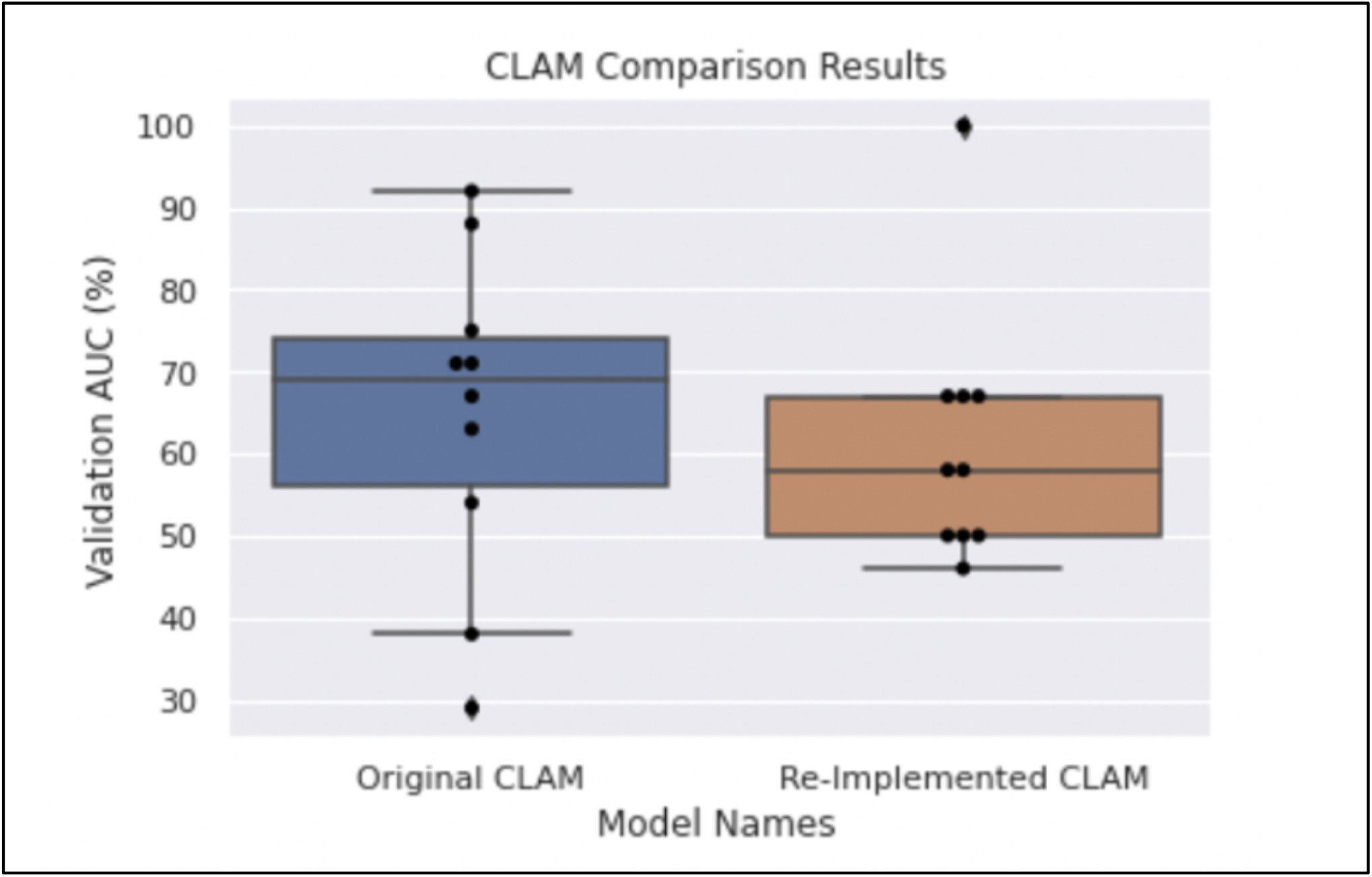
CLAM comparison box plot for the TCGA dataset. Each black dot represents the validation classification AUC scores from each of the 10-fold cross-validation sets. Left). Box plot for the original Pytorch-Version CLAM; Right). Box plot for the Tensorflow-Version re-implemented CLAM.

### 3.2. Model Performance Comparison on the BACH Dataset

The validation accuracies of both the non-specialized classification models using DenseNet201, InceptionV3, One-Shot Learning, ResNet152, and VGG19 with each of their corresponding image preprocessing applied and optimized hyperparameter configurations, and the digital pathology-specialized CLAM models with seven different combinations of image preprocessing and hyperparameter configurations are listed in **Table 1**. Among the results returned by the experiments, the DenseNet201 model (indexed as D1 in **Supplementary Table 1)**, was the best performing model in classifying normal versus tumor breast tissues from the BACH dataset with a 98.16% validation accuracy. The optimal image standardization and hyperparameter configurations included the Adam optimizer, BinaryCrossEntropy as the loss function, learning rate=1E-05, batch size=20, and number of epochs=20.

**Table 1.** Results table including the validation accuracy of the non-specialized and digital pathology-specialized model architectures with different hyperparameter configurations.

| Model Index | Model Name | Model Category | Validation Accuracy<br>(mean ± std) |
| --- | --- | --- | --- |
| D1 | DenseNet201 | Non-Specialized | 98.61% ± 1.13% |
| R1 | ResNet152 | Non-Specialized | 97.08% ± 0.78% |
| I1 | InceptionV3 | Non-Specialized | 95.29% ± 0.23% |
| V1 | VGG19 | Non-Specialized | 89.48% ± 0.66% |
| O1 | One-Shot Learning | Non-Specialized | 82.40% ± 9.31% |
| C1 | CLAM | DP- Specialized | 60.00% ± 6.80% |
| C2 | CLAM | DP- Specialized | 64.00% ± 4.87% |
| C3 | CLAM | DP- Specialized | 64.00% ± 9.74% |
| C4 | CLAM | DP- Specialized | 63.00% ± 3.54% |
| C5 | CLAM | DP- Specialized | 50.00% ± 0.00% |
| C6 | CLAM | DP- Specialized | 51.00% ± 1.73% |
| C7 | CLAM | DP- Specialized | 56.00% ± 5.12% |

### 3.3. Hyperparameter Tuning in Breast Cancer Classification Model Development

Hyperparameter tuning is critical to boost the classification performance, in addition to the model architecture. The results shown in **Table 1** indicated that with the optimal hyperparameter configurations, the non-specialized image classifiers, including the DenseNet201, ResNet152, InceptionV3, VGG19 with the transfer learning approach, and the One-Shot Learning approach, could outperform the digital pathology specialized model architecture, CLAM. This suggests that computational pathologists may need to focus more on hyperparameter tuning, rather than designing more complex digital pathology specialized model architectures. The learning rate has a higher impact on both the non-specialized and digital pathology-specialized classifiers performance compared with the rest of the hyperparameters (i.e., options of optimizers and loss functions, dropout rate, batch size, and number of epochs), and thus should be the first parameter to augment when optimizing models.

In addition to manual hyperparameter tuning, the automated hyperparameter searching algorithm is another option in selecting the optimal hyperparameter configurations. Therefore, future work could adopt automated hyperparameter tuning, which could improve the efficiency of the process to identify the optimal hyperparameter configurations.

### 3.4. Impacts of Dataset Differences on CLAM Performance

Dataset difference could affect the classification model performance, in addition to model architecture, and hyperparameter configurations. The unique architecture of the CLAM model led to the performance gap of CLAM on the BACH and TCGA dataset. CLAM is an attention-based multiple-instance learning image classifier, the attention module in the CLAM architecture first assigns attention scores to each of the patches from a certain WSI, then use the top- and least-k patches sorted from their corresponding attention scores as the positive- and negative-examples of the slide-level label. Since all patches in the BACH dataset are only informative tissue, each contributes equally to the slide-level label. This deviation violates the expectation of the CLAM model, that weights informative and non-informative patches - inherently assuming that some of the images are non-informative. Therefore, CLAM should only be used when slides contain both informative and non-informative features.

DenseNet201, a non-specialized image classification model, had the highest validation accuracy (98.16%) in the breast cancer classification in this cohort. This study also indicates the impacts of hyperparameter configurations, and dataset differences, have a significant impact on image classification model performance. This suggests that digital pathology researchers must be careful to understand the strengths and limitations of choosing a model that is suited to the task at hand.

## Data Availability

All data produced in the present study are available upon reasonable request to the authors

## Acknowledgements

All authors listed have the equal contribution of this study. QG and NP implemented the model architectures and conducted the experiments. SNH and QG designed the experiment. SNH supervised the study. This study is partially supported by the University of Minnesota Graduate School Doctoral Dissertation Fellowship for the year of 2022-2023. The results shown in **Figure 2** are in whole based upon data generated by the TCGA Research Network: https://www.cancer.giv/tcga. Prof. Sheryl L. Holt (University of Minnesota) and Kristin E. Cardiel Nunez (Mayo Clinic Alix School of Medicine) provided the support of English language editing and review services.

## Declaration of Generative AI in Scientific Writing

All authors declare that the ChatGPT was used to assist language editing in the writing process. The use of ChatGPT was done with the authors oversight, control, and was carefully reviewed and edited by the authors.

