## Supplementary Table 1 for "Deep Learning in Automating Breast Cancer Diagnosis from Microscopy Images"

**Supplementary Table 1.** *Data Preprocessing and Hyperparameter Configurations Summary Table for the Digital Pathology-Specialized (CLAM) and Non-Specialized Image Classifiers (i.e., DenseNet201, InceptionV3, One-Shot Learning, ResNet152, and VGG19)*.

| **Model Index** | **Model Name** | **Model Category** | **Data Preprocessing Technique** | **Optimizer Option** | **Loss Function Option** | **Learning Rate** | **Dropout Rate** | **Batch Size** | **Number of Epochs** |
| --- | --- | --- | --- | --- | --- | --- | --- | --- | --- |
| C1 | CLAM | DP-Specialized | Image Standardization, Image Feature Normalization | Adam | BinaryCrossEntropy | 2E-04 | 0.5 | 48 | 20 |
| C2 | CLAM | DP-Specialized | Image Standardization | Adam | BinaryCrossEntropy | 1E-04 | 0.25 | 48 | 20 |
| C3 | CLAM | DP-Specialized | Image Standardization | Adam | BinaryCrossEntropy | 1E-05 | 0.25 | 48 | 50 |
| C4 | CLAM | DP-Specialized | Image Standardization | Adam | BinaryCrossEntropy | 5E-05 | 0.25 | 48 | 20 |
| C5 | CLAM | DP-Specialized | Image Standardization | Adam | Hinge | 5E-05 | 0.25 | 48 | 20 |
| C6 | CLAM | DP-Specialized | Image Standardization | SGD | CosineSimilarity | 2E-03 | 0.25 | 48 | 20 |
| C7 | CLAM | DP-Specialized | Image Standardization | SGD | BinaryCrossEntropy | 3E-03 | 0.25 | 48 | 20 |
| D1 | DenseNet201 | Non-Specialized | Image Standardization | Adam | BinaryCrossEntropy | 1E-05 | 0.25 | 20 | 16 |
| I1 | InceptionV3 | Non-Specialized | Image Standardization | Adam | BinaryCrossEntropy | 1E-05 | NA | 20 | 7 |
| O1 | One-Shot Learning | Non-Specialized | Image Standardization | Adam | BinaryCrossEntropy | 1E-04 | NA | 32 | 5 |
| R1 | ResNet152 | Non-Specialized | Image Standardization | Adam | BinaryCrossEntropy | 1E-05 | NA | 20 | 5 |
| V1 | VGG19 | Non-Specialized | Image Standardization | Adam | BinaryCrossEntropy | 1E-05 | NA | 20 | 6 |
